# Intelligent Decision Support System Facilitating Early Detection of Cardiovascular Disease

**DOI:** 10.1101/2025.10.07.25337464

**Authors:** Dhruva Nandi, Karuna Nidhi Kaur, Lita Mohanan, Krithika Ramachandran, Muthukumar, Amaresh Ganesan, Priya Ranjan, Sundhar Mohandas, K. M. Ramkumar, Rajeeve Kumar, Abraham Oomman, Jayanthi Swaminathan, Hilda Solomon, Rinku Dahiya, V.E. Dhandapani, A.H. Sruthi Anil Kumar, Harpreet Singh, Rajiv Janardhanan

**Affiliations:** SRM Medical College Hospital and Research Centre, SRM IST, Kattankulathur, Tamil Nadu; Central Leprosy Division, MoHFW, New Delhi, India; Faculty of Social Sciences, University of Wollongong, Australia; Centre for High Impact Neuroscience and Translational Applications (CHINTA), TCG Crest, Kolkata, India; SSN College of Engineering, Chennai, India; How I met your Tech, Bengaluru, Karnataka; Buddh World University, Vaishali, Bihar India; Multidisciplinary Research Unit, AIIMS, Gorakhpur, Uttar Pradesh; Department of Biotechnology, SRM IST, Kattankulathur, Tamil Nadu; Indraprastha Apollo Hospital, New Delhi; Apollo Hospital, Chennai, Tamil Nadu; Apollo Hospitals Educational and Research Foundation, Apollo Hospital, Chennai, Tamil Nadu; Apollo Hospitals Educational and Research Foundation, Apollo Hospital, New Delhi; Sri Kalpana Heart Care, Chennai, Tamil Nadu; Bioinformatics Centre, Indian Council of Medical Research, New Delhi

## Abstract

**Background:** Cardiovascular disease (CVD) remains the primary cause of mortality worldwide, with higher fatality rates in India. Multi-modal diagnostics integrating electrocardiogram (ECG) analysis, cardiac biomarkers, and region-specific insights can enhance early detection and clinical triage.

**Methods:** In this cross-sectional study, ECGs along with clinico-epidemiological data were collected from two regions-North and South Indian cohorts. ECGs were clinically annotated and were analyzed using Earth Movers Distance (EMD) and Support Vector Machine (SVM). A 2 ml blood sample from both cohorts was also collected for molecular analysis. Age and sex matched serum samples were curated to detect four novel cardiac biomarkers. ROC curve analysis was used to assess the relationship between the biomarker index and the probability of ECG abnormality. GraphPad 10.5 and Stata version 17.0 was used for statistical analysis.

**Results:** A total of 774 clinically annotated ECGs were collected (498 from North and 276 from South Indian cohort). For the molecular analysis, 34 serum samples were curated the North Indian cohort and 54 from the South Indian cohort. The proposed SVM algorithm reported combined accuracy of 90% for non-biomarker analyzed ECGs and 94% accuracy for biomarker analyzed ECGs. South Indian cohort with abnormal ECGs reported significantly higher GDF-15 level (1145.8 ± 476.7 pg/ml) (p<0.001) as compared to North Indian cohort. GDF-15 also exhibited potential prediction ability for detecting abnormal ECG findings (AUC-0.8853).

**Conclusion:** Our study demonstrates GDF-15 as a potential biomarker for detecting region-specific abnormal ECG patterns. The proposed multimodal platform’s high diagnostic accuracy reinforces the efficacy of the artificial intelligence-driven approach in early screening and triaging CVD cases across a diverse Indian population.

**Funding:** This study was funded by the Indian Council of Medical Research (ICMR) under its Ad-hoc Research Grant Scheme (Proposal ID: 2020-3500).

## Introduction

According to the World Health Organization (WHO), cardiovascular disease (CVD) is the leading cause of death globally, impacting an estimated 17.9 million people each year (1). Case fatalities from CVD appear to be significantly greater in lower-middle income nations, including India, than in middle- and high-income countries (2, 3). Over the previous few decades, the prevalence of coronary heart disease (CHD) in India has ranged from 1.6% to 7.4% in rural areas and 1% to 13.2% in urban areas (4). Despite wide variations in risk factor prevalence, CVD-related death rates in India have resulted in an epidemiological shift from infectious to non-communicable diseases during the last two decades (5, 6). Lack of resources for triaging or stratifying patients based on the severity of their ailment results in a prolonged treatment period, reducing the patient’s prognosis. Additionally, the paucity of cardiologists (7) and the growing number of people suffering from heart disease, along with population-level changes in urban and rural areas, such as shifts in diet, lifestyle, and environmental factors, have worsened the situation (8). Although an electrocardiogram (ECG) signal can help predict various CV illnesses and provide valuable insights into regional variations and susceptibilities, (9) however, to understand the impact of differences in genetics, environment, and lifestyle, it is imperative to develop automated and accurate procedures for ECG-based diagnosis. Particularly when comparing two epidemiologically distinct regions of India, i.e. Northern and Southern regions of India, in terms of CVD manifestation and progression, this will not only complement cardiologists in improving the diagnosis, but it will also provide triaging, resulting in enhanced patient clinical outcomes in resource-limited settings.

The preliminary diagnosis of CVDs is decided by four factors: (i) clinical history, (ii) physical examination, (iii) ECG variables, and (iv) detection of cardiac biomarkers. Traditional risk markers, such as cholesterol (10), have facilitated the development of more complex indicators, including troponin levels, lipoprotein a (11), B-type natriuretic peptides (BNPs) (12), and C-reactive protein (CRP). Also, Soluble ST2 (sST2), growth differentiation factor (GDF)-15, and high sensitive troponin T (hsTnT) are promising novel biomarkers for Heart Failure (HF) evaluation.

Appropriate assessment of the incremental yield of a biomarker for predicting CVD risk requires appropriate evaluation of its calibration potential (13). This necessitates the development of biomarker-based triaging, which needs to be corroborated by the epidemiological and demographical characteristics of the subjects. Additionally, recent developments in genomics and bioinformatics have greatly aided in better understanding the complex nature of CVD etiology (14). However, developing an artificial intelligence-based computational platform predictive engine that utilizes genetic biomarkers to assess the risk of CVD in patients is still in its early stages (15–17). This forms a rationale for the development of robust Multi-Modal diagnostic approaches for the Indian setting, especially for the rural and semi-urban milieu, which will not only provide region-specific diagnostic insights but will also provide real-time allocation of clinical resources in community settings with varied epidemiological differences. Hence, our study aims to i) Develop and validate of an Intelligent Decision Support System for classifying ECG traces to detect CVD anomalies in both tertiary care settings and extended community and ii) Integrate and validate this multi-modal tool in clinical practice involving automated processing of anonymised ECG-traces along with conventional molecular biomarkers of CVD forming the rationale for effective triage methods to prioritise clinical intervention of the patients with severity of the diseases. The combined factors will play a key role in improving the diagnosis, upgrading the traditional diagnostic and risk stratification to not only provide prioritized clinical intervention but also identify the lurking precursor of CVDs at the community level.

## Methodology

### Data description

This was a cross-sectional study conducted in two cohorts, namely, the Northern and Southern Indian populations, who attended the cardiovascular unit of a tertiary healthcare centre for the collection, characterisation, and analysis of ECGs and molecular data. Respondents (also mentioned as subjects/participants/ patients) from all age groups who had given written informed consent to be a part of the study prior to the inclusion in the study were only considered. For the northern region (the selected region was New Delhi), the informed consent form was made in Hindi and English; for the southern region, the informed consent form was made in Tamil and English (the selected region was Tamil Nadu). Those who did not give informed consent for the study were excluded from the study. Purposive sampling was used for the selection of the respondents for both regions. As one of our study objectives was based on the comparative assessment of biomarkers for CVDs in the North and South regions of Indian demographics, subjects recruited in the Northern region with South-Indian ethnicities were excluded from the study. Similarly, subjects recruited in the Southern region with North-Indian ethnicities were excluded from the study. No sampling weights or adjustments were applied during analysis. A proper Case Report Form (CRF) was completed after taking each patient’s detailed medical history and socio-demographic profile. The blood investigation and ECG reports of each patient were also included in the CRF. The routine blood investigations and lipid profiling done as a standard protocol for the patient during their visit in the cardiac OPD have been recorded from the respective patient’s medical data. After completing every

CRF, the assigned cardiologist reviewed them and diagnosed each ECG as normal or abnormal by indicating the elevated pointers in the ECG graph. After completing each patient’s CRF form, the data were extracted from the CRF forms and entered into the Excel sheets systematically. To maintain confidentiality, each patient has been given a unique identification number separated for the North and South Indian cohorts to prevent bias and challenges associated with the classification of the patients.

### Data Collection

774 ECGs (532 normal annotated ECGs and 242 abnormal annotated ECGs) were collected from both North and South Indian cohorts. The sample of 774 electrocardiograms (ECGs) included in this study was derived based on operational feasibility, availability of patient data, and the aim to ensure adequate representation of both normal and abnormal ECG patterns across two demographically and geographically distinct populations — North and South India. Complete-case analysis was performed, and participants with missing clinical data were excluded from relevant analyses. No imputation methods were applied, and missing data were not systematically modelled. 498 ECGs (422 normal annotated ECGs + 76 abnormal annotated ECGs) were collected from North India, and 276 ECGs (89 normal annotated ECGs + 187 abnormal annotated ECGs) were collected from South India (Table 1). The blood was also collected for the molecular analyses. A 2 ml blood sample was collected for molecular analysis from 34 participants from the North Indian cohort and 54 participants from the South Indian cohort. No additional data were collected beyond the 774 ECGs. The participants selected for biomarker analysis were purposefully selected from within this original cohort of 774 individuals who had provided ECG data. The blood was centrifuged, and 1 ml of the serum was extracted and frozen for further molecular analyses. The data variability and the limited number of blood samples for molecular analysis from the North and South Indian cohorts were due to ethical and operational challenges.

**Table 1:** The detailed description of the dataset. Table 1 describes the clinically annotated ECG dataset collected and segregated from the North and South Indian cohorts, each representing a unique individual (biological replicates). No technical replicates were included. The data are categorized into two main groups: (1) Non-biomarker analyzed ECG data from North and South India (n= 774), and (2) Biomarker analyzed ECG data from North and South India (n = 88). In each category, the data are further stratified into North Indian, South Indian, and combined cohorts. The number of normal and abnormal ECGs for each subgroup has also been detailed. (No additional data were collected beyond the 774 ECGs. The participants selected for biomarker analysis were purposefully selected from within this original cohort of 774 individuals who had provided ECG data).

| **Categories** | **Utilized**<br>**ECGs**<br>**(n)** | **Normal**<br>**ECGs**<br>**(n)** | **Abnormal**<br>**ECGs**<br>**(n)** |
| --- | --- | --- | --- |
| **Non-biomarker analyzed ECG data from North and South India** |  |  |  |
| Combined data (North + South) | 774 | 532 | 242 |
| North Indian data | 498 | 422 | 76 |
| South Indian data | 276 | 89 | 187 |
| **Biomarker analyzed ECG data from North and South India** |  |  |  |
| Combined data (North + South) | 88 | 38 | 50 |
| North Indian data | 34 | 20 | 14 |
| South Indian data | 54 | 18 | 36 |

### Computational analyses

Our research centres around the application of computational techniques combined with molecular data with an aim to interpret ECG signals from two regions of the Indian sub-continent. We specifically focused on the binary classification of these signals into ’normal’ and ’abnormal’ categories. Two primary computational tools were employed: Earth Movers Distance ‘EMD’ for feature extraction from ECG signals, and Support Vector Machine (SVM) for classification.

- Earth Mover’s Distance (EMD) in ECG Feature Extraction:

EMD, in the context of ECG signals, measures the least amount of work needed to transform one ECG signal into another, which can be seen as a form of distance metric between two signals. The potency of EMD lies in its ability to compare signals of differing lengths and capture temporal and morphological differences, making it especially suitable for ECG signals that can vary considerably among individuals.

Given two ECG signals P and Q, the EMD is defined as:

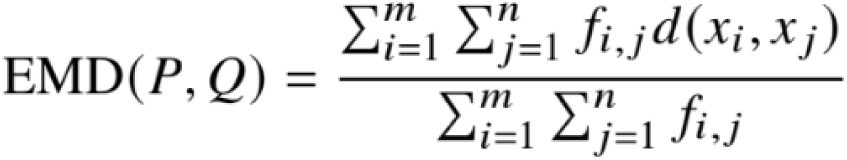

Where:

- fi, j denotes the flow between pi and q j
- d (xi, xj) is a metric (like Euclidean) that measures the distance between ECG sample points xi and x j

### Support Vector Machines (SVM) for ECG Classification

SVM is a supervised machine learning model suited for binary classification tasks. Given the features extracted from ECG signals using EMD, SVM aims to find a hyper plane that best discriminates between the ’normal’ and ’abnormal’ categories. Let’s denote the extracted ECG feature vector as x and its corresponding label (either ’normal’ or ’abnormal’) as y. The SVM’s optimisation task can be formulated as:

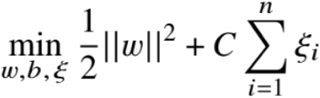

Subject to:

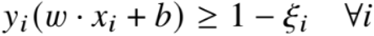

Where:

- w represents the weight vector
- b is the bias term
- îi are the slack variables accounting for misclassification
- C is a regularization parameter to balance margin maximization and error minimization

The combination of EMD feature extraction and SVM classification allows for a robust approach to discerning intricate variations in ECG signals, potentially identifying regional disparities in cardiovascular health between North and South India.

Accurate classification of instances within our dataset is symbolised by true positives (TP) and true negatives (TN). A TP illustrates a precise prediction of the positive class, while a TN reflects an accurate prediction of the negative class. Conversely, a false positive (FP) occurs when a negative instance is incorrectly identified as positive, and a false negative (FN) happens when the model inaccurately labels a positive instance as negative. Within the context of our analyses, a model that accumulates a higher number of FNs is perceived as less dependable since it overlooks vital positive class identifications. Conversely, a model with a higher TP count is regarded positively, indicating its capability to identify critical classifications accurately. The confusion matrix concisely encapsulates the allocation of TP, TN, FP, and FN in a classification context. From this matrix, we extract the subsequent performance metrics:

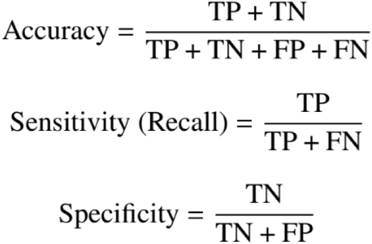

Accuracy is a dependable metric when the dataset demonstrates a balanced class distribution. Regarding model capabilities, sensitivity or recall measures the model’s adeptness in identifying true positives. Concurrently, specificity assesses the model’s ability to identify true negatives accurately.

### Molecular data analyses

The age and sex-matched serum samples from both the North and South Indian cohorts were curated for clinical and molecular analyses to detect novel biomarkers, such as soluble suppression of tumorigenicity-2 (sST2, also denoted as ST2), Heart-type Fatty Acid Binding Protein (H-FABP), Growth Differentiation Factor – 15 (GDF-15), and High-sensitive Troponin T (hs-Trop T), for predicting impending cardiovascular events at both tertiary care centres and the extended community.

The Human-specific Enzyme-Linked Immunosorbent Assay (ELISA) kits were used to measure GDF-15, H-FABP, Cardiac Troponin-T, and ST-2 levels. 50μl of plasma samples were loaded into each well of the ELISA plate. 50μl of appropriate antibody cocktail was then added to the wells after which the plate was sealed and incubated for 60 minutes at room temperature. After the incubation period, the contents of the wells were decanted, and the wells were washed three times, which was followed by the addition of 100μl of Tetramethylbenzidine (TMB) Development. After 10-minute incubation in dark, the color reaction was halted by adding 100μl stop solution without discarding the contents of the well. The absorbance was quantified using plate reader (INFINITE 200, Tecan Life Sciences, USA) at a wavelength of 450nm. The levels of study markers in plasma samples were denoted in pg/mL.

### Statistical analyses

The clinico-epidemiological data of the North and South Indian cohort were collected in the CRFs with the unique identification number. The collected data was manually entered into the Excel sheet for further analyses. The data from both sites were reviewed and cleaned, and descriptive analyses were applied. The data was demonstrated based on frequency and percentages. To study the association between four cardiac biomarkers (Cardiac Troponin-T, Human FABP, GDF-15 and Human ST-2) with the molecular analyzed normal and abnormal ECGs from both North and South Indian cohorts, Mann-Whitney unpaired t-test, one-way ANOVA and Turkey’s test was performed using GraphPad 10.5. Additionally, the study looked at four cardiac-related biomarkers, namely GDF-15, ST2, H-FABP, and hs-Trop T, and their associations with ECG findings. Each biomarker was standardized using the z-scores to remove the scale differences and to ensure comparability. The biomarkers were then directionally aligned so that higher standardized values would indicate increased biological risk. Then, a composite index called the standardized biomarker index was constructed by averaging the standardized scores of these biomarkers. The standardized biomarker index was treated as a continuous predictor in regression models. Stata version 17 was used to perform the analysis.

A predictive probability plot was generated to visualize the relationship between the biomarker index and the probability of ECG abnormality. Furthermore, ROC curve analysis of the biomarkers was conducted to evaluate their ability to discriminate between normal and abnormal ECG status. All analyses were conducted using Stata version 17.0.

### Ethical clearance

This study was conducted in accordance with the Declaration of Helsinki. Ethical approval was obtained from the institutional ethics committee, and informed consent was secured from all human participants prior to their inclusion in the study, as well as regarding the blood sample used.

### · Ethical clearance for the North Indian cohort-

Ethical clearance was received from the Ethics Committee BioMedical Research (BMR) [PROTOCOL 2020-3500 Version 2.0, 12-08-2021 (Ref. IEC: IAH-BMR-022/05-21)].

### · Ethical clearance for the South Indian cohort-

After Interim modifications, ethical clearance was received from the Ethics Committee Bio Medical Research (BMR) [PROTOCOL 2020-3500 Version 4.0, 13-12-2021 (Ref. IEC: AMH-C-S-025/06-2021)].

### · Ethical clearance for the site of experiment-

An ethical approval was taken from the institutional ethics committee (Ref. No. IBSC/AUUP/2020-1 /6).

## Result

After classifying the ECG images into normal and abnormal categories using EMD, the accuracy of the computational platform was assessed by SVM algorithm (Table 2). In category 1, we have reported 90% overall accuracy at the cost of intense computation investment after combining non-molecular analyzed ECGs of both north and south Indian cohorts. In category 2, the overall accuracy of the biomarker analyzed ECG data from North and South Indian cohort was 94%. We have curated serum samples from the included participants from both the cohorts for clinical and molecular analyses to detect novel biomarkers including sST2, H-FABP, GDF-15, hs-Trop T for predicting impending cardiovascular events. A total of 34 curated serum samples were collected from the North Indian cohort out of which 20 had clinically annotated normal ECGs (62%) and 14 abnormal ECGs (38%) (Figure 1) (Supplementary figure 1). A total of 54 curated serum samples were collected from the South Indian cohort, out of which 18 documented clinically annotated normal ECGs (33.3%) and 36 as abnormal ECGs (66.7%) (Figure 2) (Supplementary figure 2). All the samples were age and sex matched. The socio-demographic profile of the participants (for the molecular analyses) from North and South Indian cohort are described in Table 3. The most common clinical symptoms reported in the North Indian cohort (for the molecular analyses) included shortness of breath (11.8%) and chest pain (8.8%), suggesting possible cardiovascular stress. Medical history indicated smaller proportions reporting conditions such as diabetes (3%), dyslipidemia (3%), and cardiovascular disease (3%) (Table 4). Majority of the participants had family histories of cardiovascular disease (22.2%) and high blood pressure (14.8%) (Supplementary Table 1).

**Figure 1.**
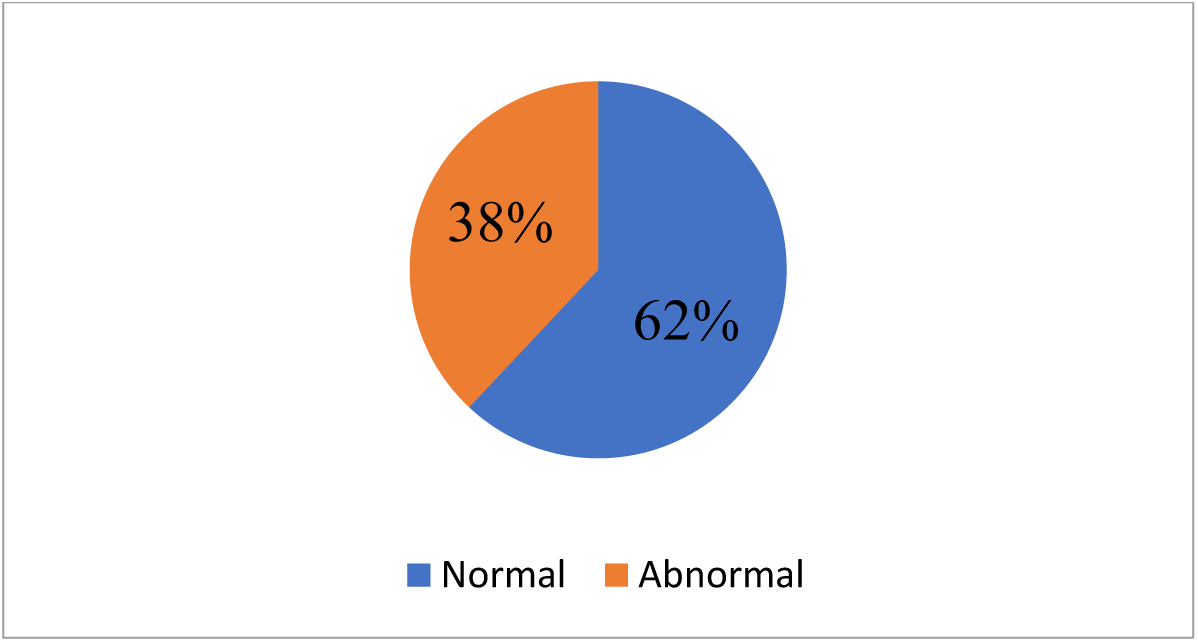
Figure depicting the electrocardiogram status of the participants (biomarker analyzed) from North Indian cohort. Figure 1 shows the pie representation of the electrocardiogram status (biomarker analyzed) of the selected 34 participants (biological replicates: n = 34; no technical replicates) from the North Indian cohort. Groups were divided based on ECG status: normal (n = 20) (62%), abnormal (n = 14) (38%).

**Figure 2.**
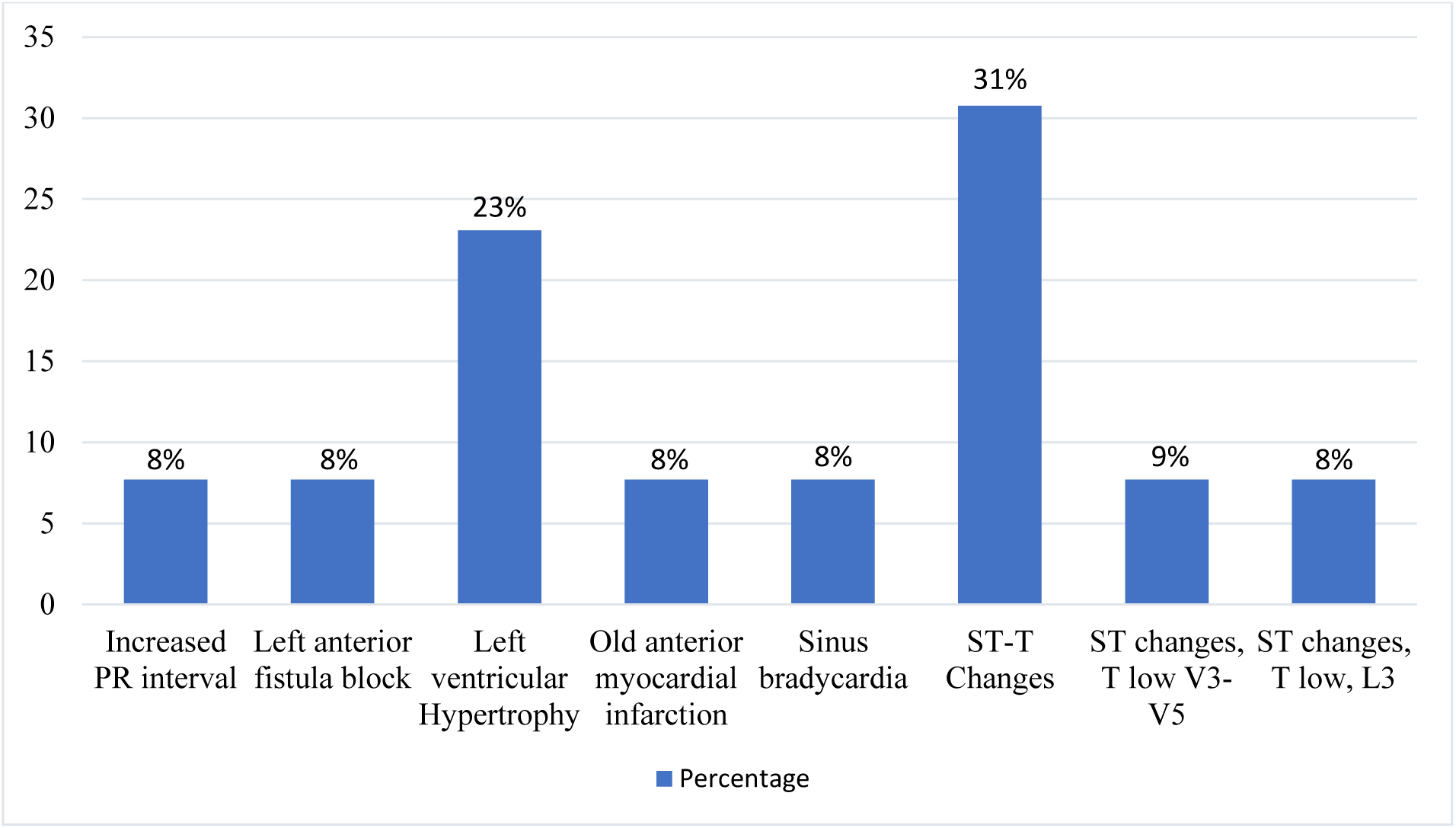
Figure describing the abnormal electrocardiogram findings of the participants (for the molecular study) from North Indian cohort. Figure 2 describes the various abnormal electrocardiogram findings (biomarker analyzed) in the 14 participants (biological replicates: n = 14; no technical replicates). 31% had abnormal ST-T changes, 23% had left ventricular hypertrophy and 8% had old anterior myocardial infarction. Combined ST changes and T low V3-V5 was reported in 9% of the participants. Also, an abnormal combination of ST changes, T low and L3 was found in 8% of the participants.

**Figure 3.**
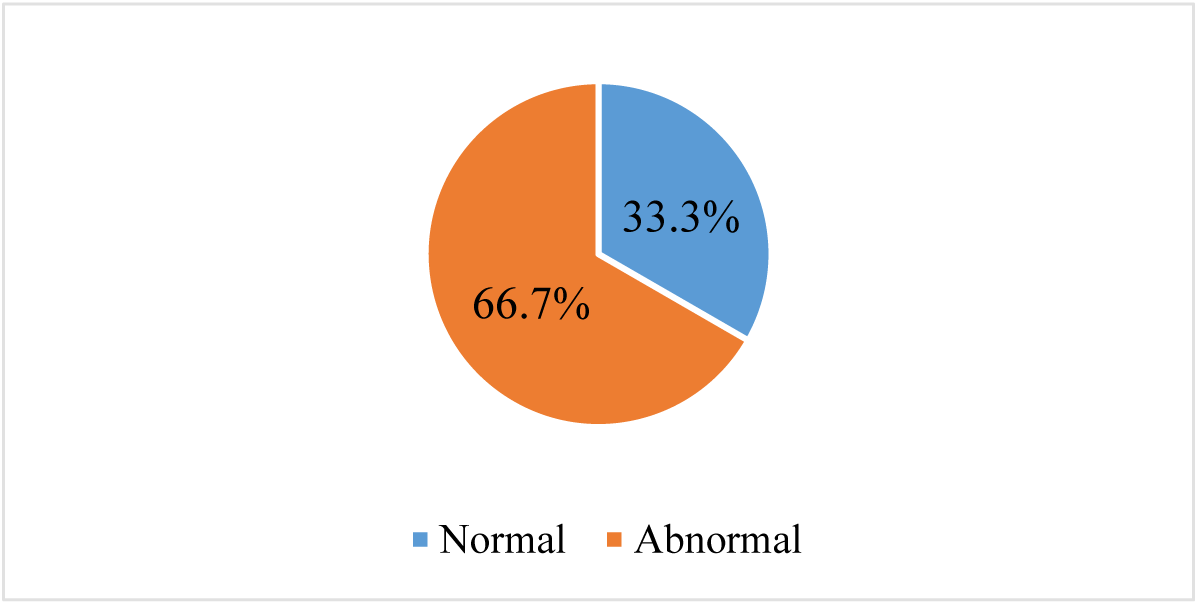
Figure depicting the electrocardiogram status of the participants (for the molecular study) from the South Indian cohort. Figure 3 shows the pie representation of the electrocardiogram status (biomarker analyzed) of the 54 participants (biological replicates: n = 54; no technical replicates). Groups were divided based on electrocardiogram status: normal (n = 18) (33.3%), abnormal (n = 36) (66.7%).

**Table 2:** Table depicting the accuracy (in %) of the SVM algorithm in classifying various categories of ECGs. This table presents the classification accuracy metrics derived from the clinically annotated ECG dataset (biological replicates; one ECG per individual, with no technical replicates). The dataset was divided into two groups: (1) Non-biomarker analyzed ECG data (n = 774) and (2) Biomarker analyzed ECG data (n = 88). Accuracy percentages are reported for normal ECGs, abnormal ECGs, and overall classification performance. Each group is further stratified by region—North Indian, South Indian, and combined (North + South) cohort— to assess geographic variability in model performance. The analysis was conducted as part of a broader research focus applying computational techniques integrated with molecular data to interpret ECG signals from two regions of the Indian sub-continent. Binary classification of ECG signals into ’normal’ and ’abnormal’ categories was performed using Earth Mover’s Distance (EMD) for feature extraction and Support Vector Machine (SVM) for classification.

| **Categories** | **Normal**<br>**accuracy**<br>**(%)** | **Abnormal**<br>**accuracy**<br>**(%)** | **Overall**<br>**accuracy**<br>**(%)** |
| --- | --- | --- | --- |
| Non-biomarker analyzed ECG data from North and South India |  |  |  |
| **Combined data (North + South)** | 94 | 72 | 90 |
| **North Indian data** | 93 | 13 | 86 |
| **South Indian data** | 72 | 91 | 90 |
| Biomarker analyzed ECG data from North and South India |  |  |  |
| **Combined data (North + South)** | 94 | 96 | 94 |
| **North Indian data** | 100 | 87 | 93 |

| South Indian data | 91 | 89 | 90 |
| --- | --- | --- | --- |

**Table 3.**
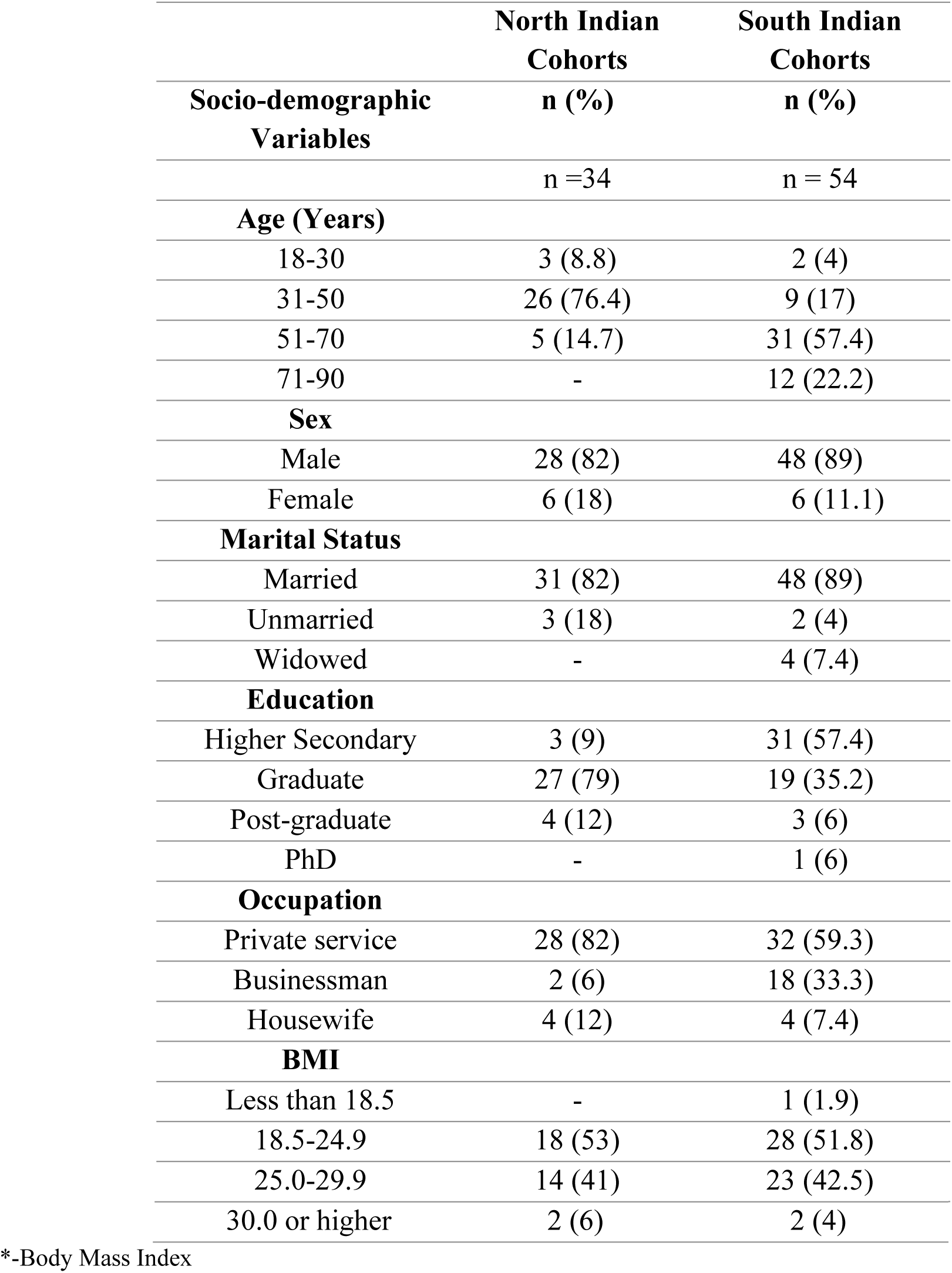
Table describing the socio-demographic profile of the participants from the North Indian and South Indian cohorts (biomarker analyzed sample) Table 3 describes the socio-demographic characteristics of participants selected for biomarker analysis from the North Indian (n = 34) and South Indian (n = 54) cohorts, totalling 88 biological replicates (each representing a unique individual). No technical replicates were included. Majority of the selected subjects from the Northern region were male (82%) and between the age of 31-50 years (76.4%).41% of the subjects were reported to be overweight falling in the BMI range of 25.0-29.9. 57.4% of the participants from the Southern region were between the age group of 51-70 years. 89% of the participants were both male and married. Majority of the participants were in the BMI range of 18.5-24.9 (51.8%). 57.4% of the participants received education at least till the higher secondary level and 59.3% are in private service jobs.

**Table 4.**
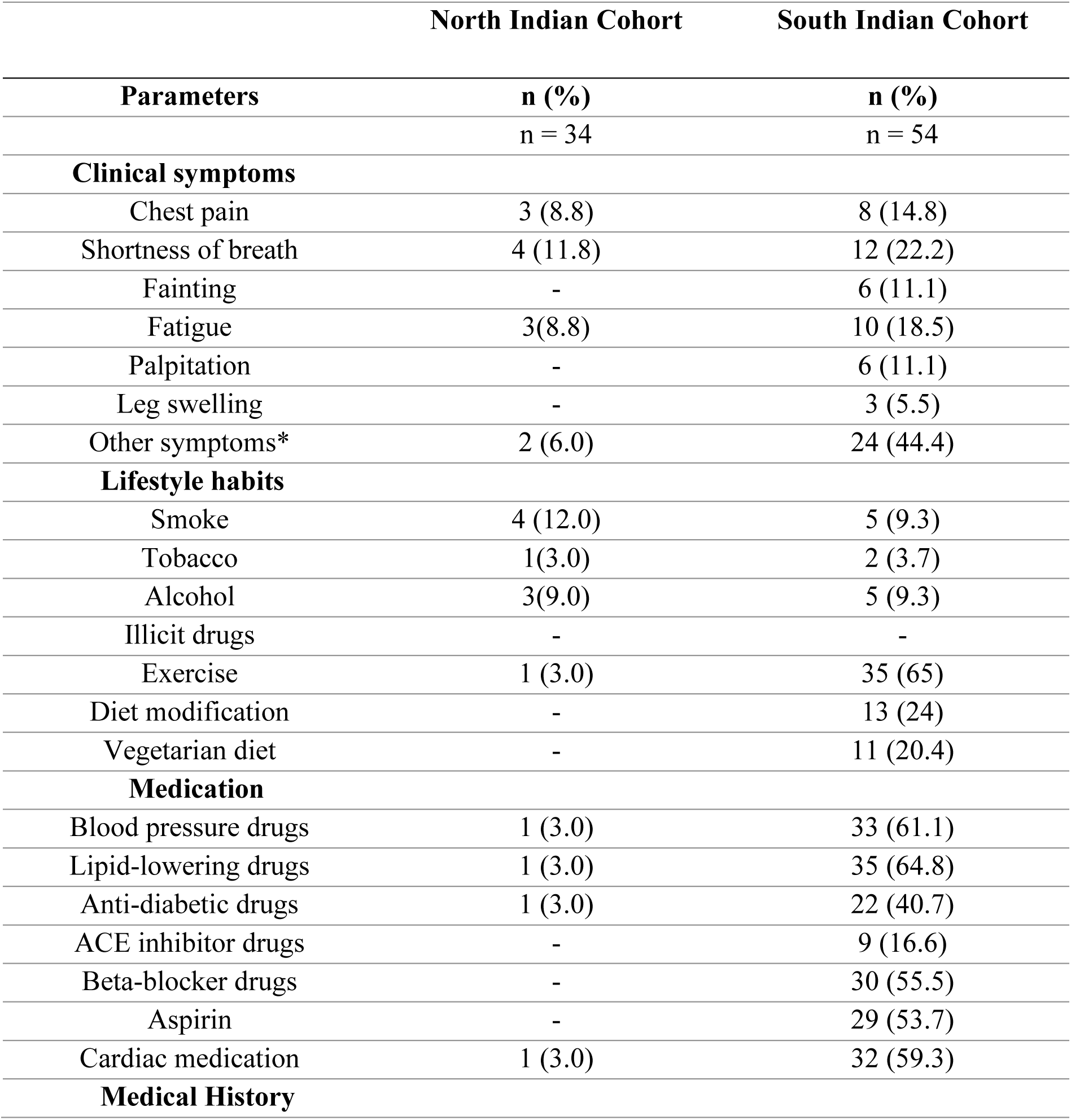
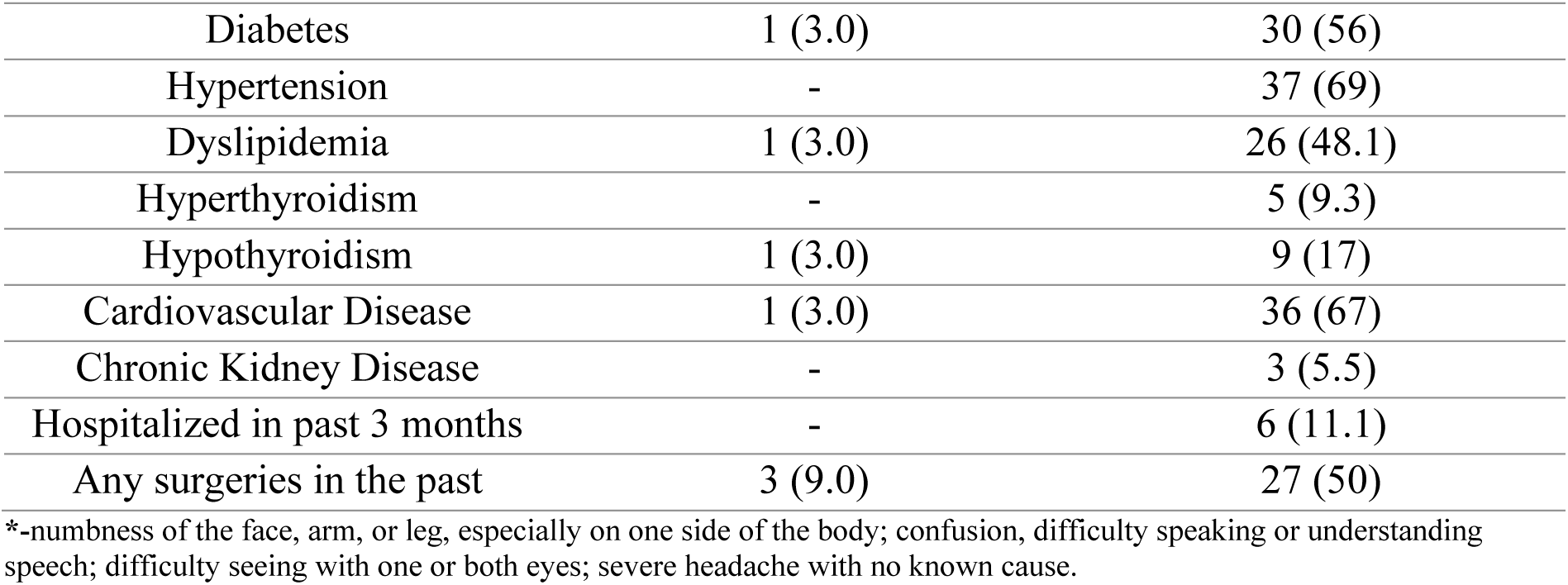
Table describing various clinical parameters of the participants from North and South India (biomarker analyzed sample) Table 4 describes the detailed medical history of the participants selected for biomarker analysis from the North Indian (n = 34) and South Indian (n = 54) cohorts, totalling 88 biological replicates (each representing a unique individual). No technical replicates were included. From the Northern cohort, 3% of the patients had a history of diabetes, 3% had Dyslipidemia, 3% had hypothyroidism, and 3% had cardiovascular disease. 9% of the patients had undergone surgeries in the past. In the southern cohort, 22.2% of the patients had shortness of breath, 18.5% reported fatigue, and 14.8% reported chest pain. Both palpitation and fainting were present in11.1% of the patients. 9.3% of the patients were smokers, 9.3% consumed alcohol and 3.7% were tobacco consumers. 65% of the patients exercised regularly. 43 patients were eating a non-vegetarian diet whereas only 11 were vegetarians. 56% of the patients had a history of diabetes, 67% had cardiovascular diseases and 69% had hypertension. 48.1% had reported dyslipidemia and 5.5% reported chronic kidney diseases. Also, 50% of the patients had undergone surgeries in the past.

|  | North Indian Cohort | South Indian Cohort |
| --- | --- | --- |
| Parameters | n (%) | n (%) |
|  | n = 34 | n = 54 |
| **Clinical symptoms** |  |  |
| Chest pain | 3 (8.8) | 8 (14.8) |
| Shortness of breath | 4 (11.8) | 12 (22.2) |
| Fainting | - | 6 (11.1) |
| Fatigue | 3(8.8) | 10 (18.5) |
| Palpitation | - | 6 (11.1) |
| Leg swelling | - | 3 (5.5) |
| Other symptoms* | 2 (6.0) | 24 (44.4) |
| **Lifestyle habits** |  |  |
| Smoke | 4 (12.0) | 5 (9.3) |
| Tobacco | 1(3.0) | 2 (3.7) |
| Alcohol | 3(9.0) | 5 (9.3) |
| Illicit drugs | - | - |
| Exercise | 1 (3.0) | 35 (65) |
| Diet modification | - | 13 (24) |
| Vegetarian diet | - | 11 (20.4) |
| **Medication** |  |  |
| Blood pressure drugs | 1 (3.0) | 33 (61.1) |
| Lipid-lowering drugs | 1 (3.0) | 35 (64.8) |
| Anti-diabetic drugs | 1 (3.0) | 22 (40.7) |
| ACE inhibitor drugs | - | 9 (16.6) |
| Beta-blocker drugs | - | 30 (55.5) |
| Aspirin | - | 29 (53.7) |
| Cardiac medication | 1 (3.0) | 32 (59.3) |
| **Medical History** |  |  |
| Diabetes | 1 (3.0) | 30 (56) |
| Hypertension | - | 37 (69) |
| Dyslipidemia | 1 (3.0) | 26 (48.1) |
| Hyperthyroidism | - | 5 (9.3) |
| Hypothyroidism | 1 (3.0) | 9 (17) |
| Cardiovascular Disease | 1 (3.0) | 36 (67) |
| Chronic Kidney Disease | - | 3 (5.5) |
| Hospitalized in past 3 months | - | 6 (11.1) |
| Any surgeries in the past | 3 (9.0) | 27 (50) |

\*-numbness of the face, arm, or leg, especially on one side of the body; confusion, difficulty speaking or understanding speech; difficulty seeing with one or both eyes; severe headache with no known cause.

Table 5 provides a detailed overview of laboratory investigations, lipid parameters, and ECG evaluation in the North Indian curated samples regarding their cardiovascular and general health condition. Abnormal ECG findings among the selected 34 participants (Figure 2) from the North Indian cohort reported ST-T changes being the most common (31%). From the curated sample data of the South Indian cohort, 24.1% of the patients had low haemoglobin level, 14.8% had low packed cell volume and 5.5% had low platelets (Table 6). Figure 4 showed 13.9% patients with old anterior myocardial infarction, indicating a history of ischemic heart disease.

**Figure 4.**
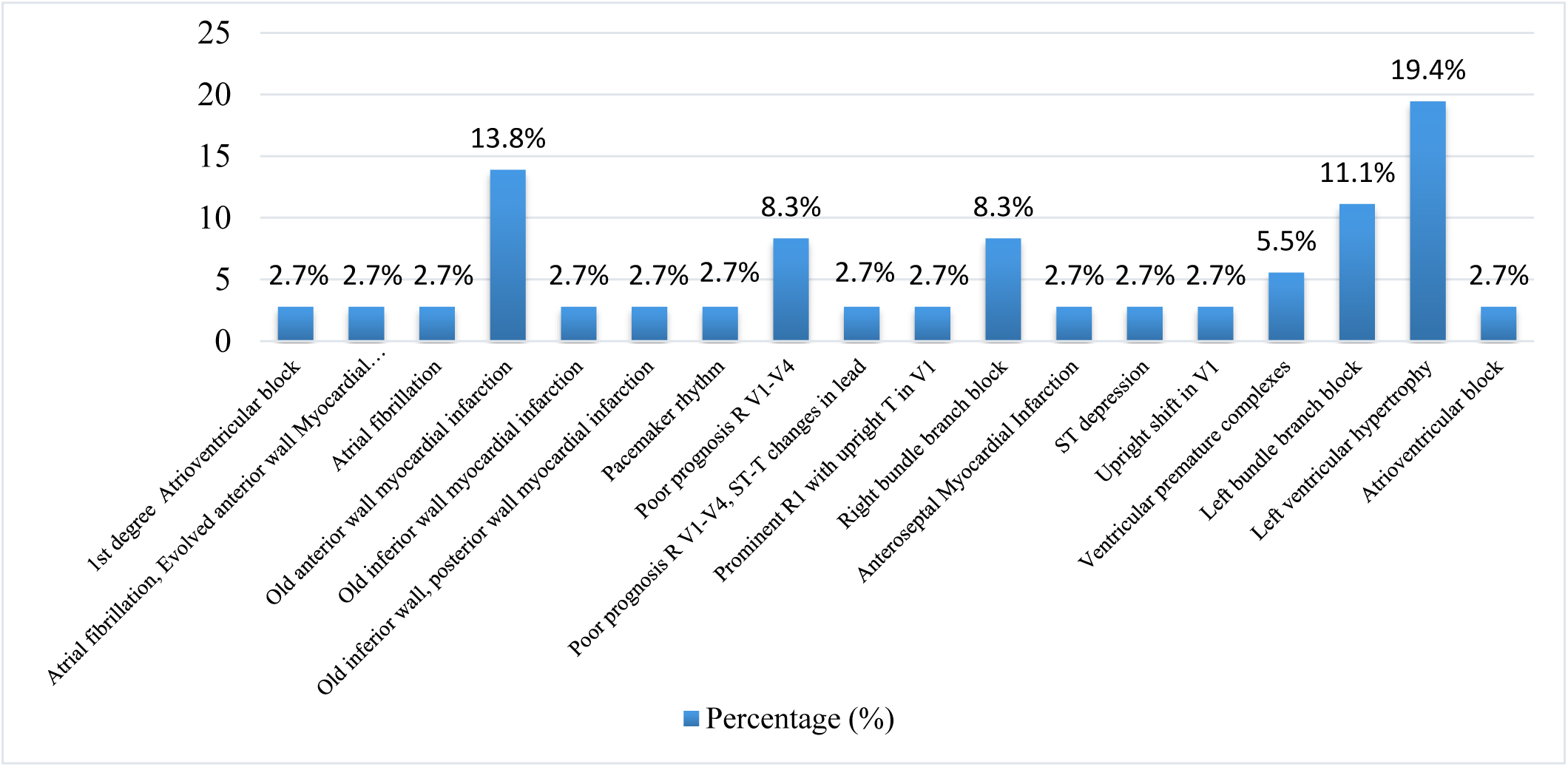
Figure depicting the abnormal electrocardiogram findings of the participants (for biomarker analysis) from the South Indian cohort. Figure 4 shows a graphical representation of the abnormal electrocardiogram findings (biomarker analyzed) of the 36 participants (biological replicates: n = 36; no technical replicates). 19.4% had left ventricular hypertrophy, 13.9% had an old anterior myocardial infarction, 11.1% had a left bundle branch block, 8.3% had a right bundle branch block, 5.5% had ventricular premature complexes and 2.7% had an old inferior myocardial infarction

**Table 5.** Table describing various laboratory investigations, lipid parameters and electrocardiogram assessment of the patients) from North Indian cohort (biomarker analyzed sample) Table 5 describes the routine laboratory investigation of the 34 patients from the North Indian cohort (each representing a unique individual). No technical replicates were included. 35% of the patients were reported to have low haemoglobin level. 18 % of the patients had low neutrophil count, 21% had low lymphocyte count and 35% had low urea. On the other hand, 50% of the patients had high RDW, 15% had high HbA1c, and 9% had high uric acid serum. 32% of the patients had high total cholesterol level and 21% had high triglyceride levels, 71% of the patients had high LDL cholesterol levels and 35% of the patients had high Cholesterol/HDL ratio. Also, 35% of the patients had low HDL cholesterol levels. Thyroid-stimulating hormone was found to be high in 15% of the patients

| Parameters | Normal | Abnormal |  | Not available |
| --- | --- | --- | --- | --- |
|  | n (%) | Low<br>n (%) | High<br>n (%) | n (%) |
| **Laboratory investigations** |  |  |  |  |
| Haemoglobin | 22 (65) | 12 (35) | - | - |
| PCV | - | - | - | 34(100) |
| RBC | 34 (100) | - | - | - |
| WBC | 34(100) | - | - | - |
| Platelet count | 30 (82) | 4(12) | - | - |
| Neutrophils | 28 (82) | 6(18) | - | - |
| Lymphocyte | 24(71) | 7(21) | 3(9) | - |
| Monocyte | 29(85) | 4(12) | 1(3) | - |
| Eosinophil | 24(71) | 5(15) | 5(15) | - |
| Basophils | 33(97) | - | - | 1(3) |
| RDW | 16(47) | 1(3) | 17(50) | - |
| ESR | 28(82) | - | 1(3) | 5(15) |
| HbA1c | 10(29) | - | 5(14) | 19(56) |
| Sodium level | 12(35) | 2(6) | - | 20(59) |
| Potassium level | 14(41) | - | - | 20(59) |
| Chloride level | 13(38) | - | 1(3) | 20(59) |
| Bicarbonate level | 1(3) | - | - | 33(97) |
| Carbon dioxide level | - | - | - | 34(100) |
| Alkaline phosphatase | 34(100) | - | - | - |
| Bilirubin serum | 22(65) | - | 2(6) | 10(29) |
| Bilirubin conjugated | 22(65) | - | 1(3) | 11(32) |
| Protein | 24(71) | - | 1(3) | 9(26) |
| Albumin | 20(59) | - | 5(15) | 9(26) |
| ALT | 24(71) | - | 2(6) | 8(24) |
| Urea | 18(53) | 12(35) | - | 4(12) |
| Creatinine | 25(74) | 1(3) | 2(6) | 6(18) |
| Uric acid serum | 24(71) | 2(6) | 3(9) | 5(15) |
| **Lipid profile** |  |  |  |  |
| Total cholesterol | 22(65) | - | 11(32) | 1(3) |
| Triglyceride | 24(71) | - | 7(21) | 3(9) |
| HDL cholesterol | 20(59) | 12(35) | - | 2(6) |
| Non-HDL cholesterol | - | - | - | 34 (100) |
| LDL cholesterol | 5 (15) | - | 24(71) | 5(15) |
| VLDL cholesterol | 29(85) | - | 1(3) | 4(12) |
| Cholesterol/HDL ratio | 20(59) | - | 12(35) | 2(6) |
| Thyroid Stimulating Hormone | 21(62) | - | 5(15) | 8(24) |
| Free T3 | 3(9) | - | - | 31(91) |
| Free T4 | 2(6) | - | - | 32(94) |
| High-sensitive Troponin I plasma | 3(9) | - | - | 31(91) |
| Creatinine kinase | - | - | - | 31(91) |
| NT-Pro BNP | - | - | - | 34(100) |
| Total cholesterol | 22(65) | - | 11(32) | 1(3) |
| Triglyceride | 24(71) | - | 7(21) | 3(9) |
| **ECG assessment** |  |  |  |  |
| Heart rate | 31(91) | 2(6) | - | 1(3) |
| QRS interval | 26(76) | 4(12) | 3(9) | 1(3) |
| QRS axis | 31(91) | - | 2(6) | 1(3) |
| PR interval | 32(94) | - | 1(3) | 1(3) |
| QTC | 33(97) | - | - | 1(3) |

PCV- Packed Cell Volume, RBC- Red Cell Volume, WBC-White Blood Cell, RDW-Red Blood Cell Distribution Width, ESR- Erythrocyte Sedimentation Rate, ALT- Alanine Aminotransferase, HDL- High Density Lipoprotein, LDL- Low Density Lipoprotein, VLDL- Very Low Density Lipoprotein, NT-Pro BNP- N-terminal pro-B-type natriuretic peptide

**Table 6.** Table describing various laboratory investigations, lipid parameters and electrocardiogram assessment of the patients from South India cohort (biomarker analyzed sample) Table 6 describes the routine laboratory investigation of the 54 patients from the South Indian cohort (each representing a unique individual). No technical replicates were included. 24.1% of the patients had low hemoglobin level, 14.8% had low packed cell volume and 5.5% had low platelets. 35.2% of the patients had high HbA1c level and 14.8% had a high erythrocyte sedimentation rate. Albumin and Creatinine levels were higher in 11.1% of the patients. 27.7% of the patients had high triglyceride levels, 22.2% had high LDL levels and 11.1% had high cholesterol/HDL ratio. Also, low HDL and non-HDL cholesterol were found to be low in 40.7% of the patients.

|  | Normal | Abnormal |  | Not available |
| --- | --- | --- | --- | --- |
| Parameters | n (%) | Low<br>n (%) | High<br>n (%) | n (%) |
| **Laboratory investigations** |  |  |  |  |
| Haemoglobin | 15 (27.7) | 13(24.1) | - | 26(48.1) |
| PCV | 17(31.5) | 8(14.8) | - | 29(53.7) |
| RBC | 20(37) | 4(7.4) | - | 30(55.6) |
| WBC | 25(46.3) | - | - | 29(53.7) |
| Platelet count | 21(38.8) | 3(5.5) | 1(2) | 29(53.7) |
| Neutrophils | 23(42.6) | - | - | 29(53.7) |
| Lymphocyte | 24(71) | 2(3.7) | 1(2) | 30(55.6) |
| Monocyte | 23(42.6) | 1(2) | - | 30(55.6) |
| Eosinophil | 22(40.7) | - | 1(2) | 31(57.4) |
| Basophils | 6(11.1) | - | - | 48(88.9) |
| RDW | 15(27.7) | - | 9(16.7) | 30(55.6) |
| ESR | 12(22.2) | - | 8(14.8) | 34(62.9) |
| HbA1c | 6(11.1) | - | 19(35.2) | 29(53.7) |
| Sodium level | 28(51.8) | 1(2) | - | 25(46.3) |
| Potassium level | 25(46.3) | - | (7.4) | 25(46.3) |
| Chloride level | 23(42.6) | - | 2(3.7) | 29(53.7) |
| Bicarbonate level | 1(1.8) | - | - | 53(98.1) |
| Carbon dioxide level | 22(40.7) | - | 1(2) | 31(57) |
| Alkaline phosphatase | 21(38.8) | 1(2) | - | 32(59) |
| Bilirubin serum | 20(37) | - | 1(2) | 33(61.1) |
| Bilirubin conjugated | 18(33.3) | - | 2(3.7) | 34(63) |
| Protein | 20(37) | 1(2) | - | 33(61.1) |
| Albumin | 13(24.1) | 2(3.7) | 6(11.1) | 33(61.1) |
| ALT | 16(29.6) | - | 6(11.1) | 32(59.3) |
| Urea | 23(42.5) | 2(3.7) | 2(3.7) | 27(50) |
| Creatinine | 23(42.5) | 1(2) | 6(11.1) | 24(44.4) |
| Uric acid serum | 21(39) | - | 3(5.5) | 30(55.6) |
| **Lipid profile** |  |  |  |  |
| Total cholesterol | 28(51.8) | - | 4(7.4) | 22(40.7) |
| Triglyceride | 18(33.3) | - | 15(27.7) | 21(39) |
| HDL cholesterol | 11(20.4) | 22(40.7) | - | 21(39) |
| Non-HDL cholesterol | - | - | - | 54(100) |
| LDL cholesterol | 22(40.7) | - | 12(22.2) | 20(37) |
| VLDL cholesterol | 1(2) | - | 1(2) | 52(96.3) |
| Cholesterol/HDL ratio | 27(50) | - | 6(11.1) | 21(39) |
| Thyroid Stimulating Hormone | 16(29.6) | - | 4(7.4) | 34(63) |
| Free T3 | - | - | - | 54(100) |
| Free T4 | - | - | - | 54(100) |
| High sensitive Troponin 1 plasma | 13(24.1) | - | - | 41(76) |
| Creatinine kinase | 13(24.1) | 2(3.7) | - | 39(72.2) |
| NT-Pro BNP | 7(13) | - | - | 47(87) |
| **Electrocardiogram assessment** |  |  |  |  |
| Heart rate | 44(81.5) | 6(11.1) | 4(7.4) | - |
| QRS interval | 38(70.4) | 5(9.3) | 11(20.4) | - |
| QRS axis | 46(85.2) | 7(13) | 1(2) | - |
| PR interval | 48(80) | - | 3(5.5) | 3(5.5) |
| QTC | 42(78) | - | 12(22.2) | - |

PCV- Packed Cell Volume, RBC- Red Cell Volume, WBC-White Blood Cell, RDW-Red Blood Cell Distribution Width , ESR- Erythrocyte Sedimentation Rate, ALT- Alanine Aminotransferase, HDL- High Density Lipoprotein, LDL- Low Density Lipoprotein, VLDL- Very Low Density Lipoprotein, NT-Pro BNP- N-terminal pro-B-type natriuretic peptide

The detailed molecular analysis of the four biomarkers, namely hs-Trop T, H-FABP, GDF-15, and sST2, taken in our study, has been discussed below.

i. **Human Cardiac Troponin-T (hs-Trop T)**

The serum level of hs-Trop T was not significantly different between the patients with normal versus abnormal ECG, when the overall study population, including both patients from North and South Indian cohort was analyzed (Figure 5a).The South Indian cohort patients with normal versus abnormal ECG also had no significant difference in hs-Trop T levels. However, the North Indian cohort patients with abnormal ECG had significantly higher hs-Trop T levels. Importantly, there were no differences between the North and South cohort patients with normal ECG indicating no effect of the socio-demographic differences on the baseline hs-Trop T levels. Next, the patients with an abnormal ECG were sub-divided into those who had a myocardial infarction (MI) event or had not had an MI. Interestingly, upon this sub-division, it was found that the hs-Trop T levels were significantly higher in patients with abnormal ECG and MI as compared to both normal and abnormal ECG patients who had not had an MI event (Figure 5b). This finding aligns with the well-established role of Cardiac Troponin-T as a biomarker for MI, but importantly, it does not increase in patients who have an abnormal ECG without an MI event.

ii. **Human Fatty Acid Binding Protein (H-FABP)**

**Figure 5.**
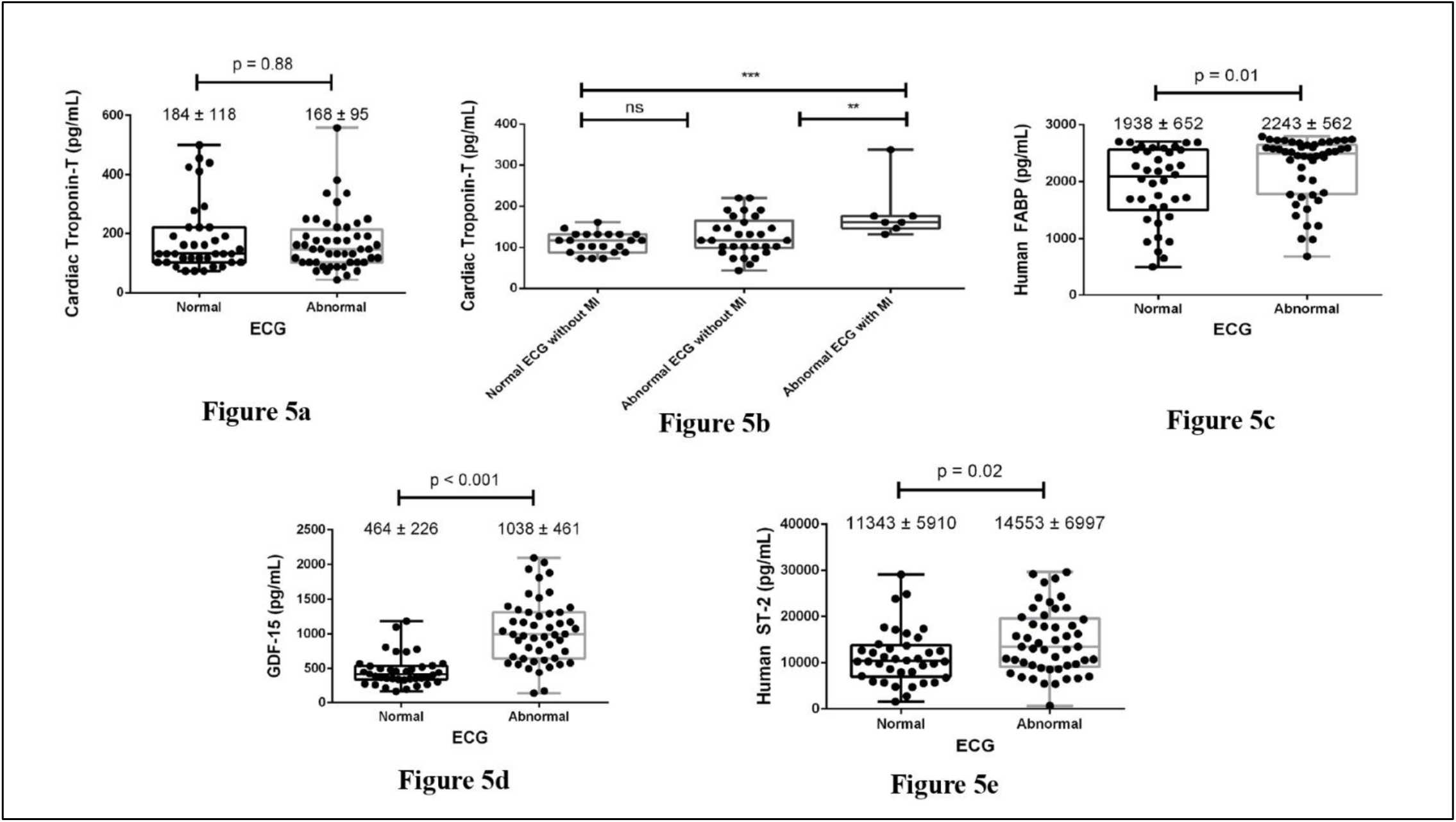
**Figure depicting the level of the four cardiac biomarkers namely hs-Trop T, H-FABP, GDF-15 and sST2 between participants with normal and abnormal ECG (biomarker analyzed) from North and South Indian cohort** Figure 5a. Cardiac Troponin-T levels between patients with biomarker analyzed normal and abnormal ECG (88 participants) (biological replicates: n = 88; no technical replicates), total normal (north + south Indian Cohort) ECGs (n = 38), abnormal (n = 50), Figure 5b. Cardiac Troponin-T (hs-Trop T) levels in biomarker analyzed normal and abnormal ECG participants with and without Myocardial Infarction in the overall study population (88 participants) (biological replicates: n = 88; no technical replicates), total normal (north + south Indian Cohort) ECGs (n = 38), abnormal (n = 50), Figure 5c. H-FABP levels between participants with biomarker analyzed normal and abnormal ECG in the overall study population (88 participants) (biological replicates: n = 88; no technical replicates), total (north + south Indian Cohort) normal ECGs (n = 38), abnormal (n = 50), Figure 5d. GDF-15 levels between participants with biomarker analyzed normal and abnormal ECG in the overall study population (88 participants) (biological replicates: n = 88; no technical replicates), total normal (north + south Indian Cohort) ECGs (n = 38), abnormal (n = 50), and Figure 5e. Human ST-2 levels between participants with biomarker analyzed normal and abnormal ECG in the overall study population (88 participants) (biological replicates: n = 88; no technical replicates), total (north + south Indian Cohort) normal ECGs (n = 38), abnormal (n = 50). Figure 5a depicts the levels of Cardiac Troponin-T in serum samples from all the patients with normal and abnormal ECGs measured by ELISA. All data are presented as mean ± standard deviation (SD). Mann-Whitney unpaired t-test was used to compare the levels of the marker between the two groups (p-value less than 0.05 is considered statistically significant). The p-value was 0.88, indicating no statistical significance. Figure 5b shows a subgroup analysis for the levels of Cardiac Troponin-T in serum samples measured by ELISA. The abnormal ECG patients have been sub-divided based on the occurrence of a myocardial infarction (MI). All data are presented as mean ±standard deviation (SD). The comparison between more than two groups was performed using one-way ANOVA, followed by multiple comparisons analyses using Tukey’s test (p-value less than 0.05 is considered statistically significant). Symbols represent: **<0.01; ***p<0.001; ns: no significance. Figure 5c shows the levels of H-FABP in serum samples from all the patients with normal and abnormal ECGs measured by ELISA. All data are presented as mean ±standard deviation (SD). Mann-Whitney unpaired t-test was used to compare the levels of the marker between the two groups (p value less than 0.05 is considered statistically significant). The p-value was 0.01, indicating statistical significance. Figure 5d documents the levels of GDF-15 in serum samples from all the patients with normal and abnormal ECGS measured by ELISA. All data are presented as mean ±standard deviation (SD). Mann-Whitney unpaired t-test was used to compare the levels of the marker between the two groups (p value less than 0.05 is considered statistically significant). The p<0.001 stated statistical significance. Figure 5e illustrates the levels of Human ST-2 in serum samples from all the patients with normal and abnormal ECGs measured by ELISA. All data are presented as mean ± standard deviation (SD). Mann-Whitney unpaired t-test was used to compare the levels of the marker between the two groups (p value less than 0.05 is considered statistically significant). The p-value was 0.02, indicating statistical significance.

The serum level of H-FABP was significantly increased in patients with abnormal ECG, when the overall study population was analyzed (Figure 5c). Interestingly, this significant difference was lost when the data was divided based on the location, which could be due to the wide range of variability among patients. Another important finding is that both in patients with normal and abnormal ECG, the South Indian cohort population had a significantly higher H-FABP level compared to the North Indian cohort population, indicating that the socio-demographic differences can influence the H-FABP levels.

iii. **Growth and Differentiation Factor-15 (GDF-15)**

In accordance to the already established role of GDF-15 as a disease biomarker, we found that there was a dramatic increase in GDF-15 levels in patients with abnormal ECG compare to those with normal ECG when we analyzed the entire study population (Figure 5d). This result held true even when the data was separated into South and North Indian cohort populations, where patients with an abnormal ECG from both centres had higher levels of serum GDF-15, making this molecule a reliable biomarker. Another critical point is that there were no differences between the North and South Indian patients with normal ECG, demonstrating no effect of the socio-demographic differences on baseline GDF-15 levels. However, among the abnormal ECG patients, it was noted that the North Indian cohort population had lower GDF-15 levels compared to the South Indian cohort population.

**iv**. Human suppression of tumorigenicity-2 (sST-2)

The serum Human ST-2 level was significantly increased in patients with abnormal ECG, when the overall study population was analyzed (Figure 5e). However, this difference was lost when the data was divided based on the location. The reason for this could be the wide range of variability among patients. Despite this variability, both in patients with normal and abnormal ECG, there was no significant difference in the ST-2 levels when the South Indian cohort population was compared to the North Indian cohort population, indicating that the socio-demographic differences do not influence the ST-2 levels. The mean and standard deviation of biomarker levels stratified by ECG status and region are presented in Table 7.

**Table 7.** Mean (Standard Deviation) of Biomarkers by ECG Status and Region. Table 7 describes Mean (standard deviation) values of individual biomarkers namely GDF-15, sST-2, H-FABP, and hs-Troponin-T stratified by region and ECG status for biomarker analysis from the North Indian (n = 34) and South Indian (n = 54) cohorts, (each representing a unique individual). No technical replicates were included. The values presented are the mean concentrations of GDF-15, sST-2, H-FABP, and hs-Troponin-T in participants with and without abnormal ECG findings. Each value represents the mean concentration (pg/mL) and standard deviation for individuals with ECGs classified as either normal or abnormal.

| Biomarkers | North Indian cohort |  | South Indian Cohort |  |
| --- | --- | --- | --- | --- |
|  | Normal ECG Mean (Standard deviation) | Abnormal ECG Mean (Standard deviation) | Normal ECG Mean (Standard deviation) | Abnormal ECG Mean (Standard deviation) |
| GDF-15 (pg/mL) | 472.8 (275.4) | 766.8 (284.3) | 455.6 (163.8) | 1145.8 (476.7) |
| sST-2 (pg/mL) | 10073.4 (5183.4) | 11768.7 (4540.0) | 12753.6 (6479.1) | 15667.3 (7534.3) |
| H-FABP (ng/mL) | 1709.0 (641.0) | 1824.0 (732.1) | 2191.4 (581.5) | 2398.1 (396.0) |
| hs-Troponin-T (ng/L) | 154.8 (88.1) | 398.1 (597.8) | 312.3 (421.2) | 230.0 (532.9) |

Upon ROC curve analyses, GDF-15 exhibited excellent discriminatory ability for predicting abnormal ECG findings, with an area under the curve (AUC) of 0.8853 (Figure 6a). After standardizing the biomarker index, integrating GDF-15, ST2, H-FABP, and Troponin-T into a composite score, the standardized biomarker index demonstrates good predictive performance with an AUC of 0.7789 (Figure 6b). Figure 6c depicts the predicted probability plot of abnormal ECGs based on the standardized biomarker index.

**Figure 6.**
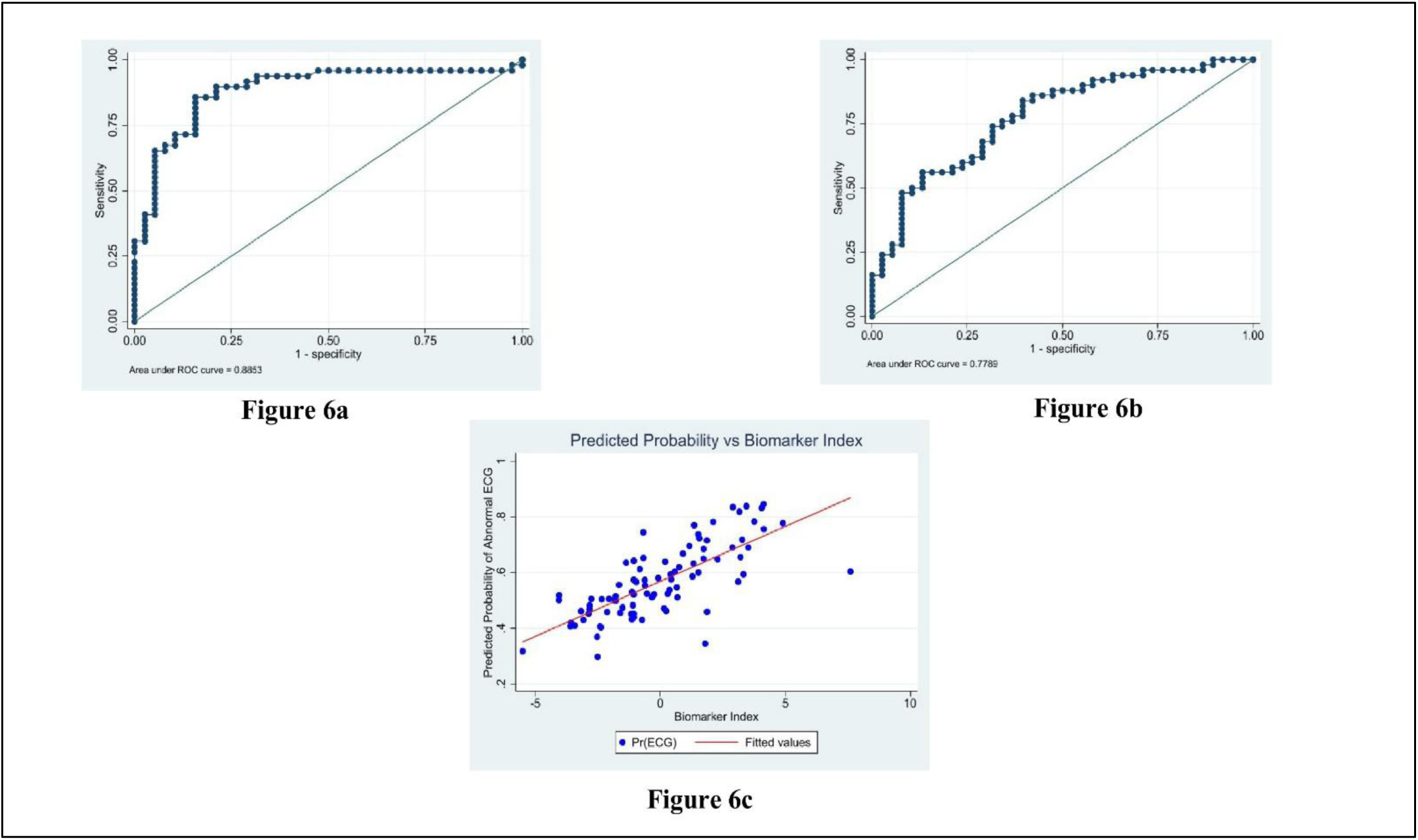
Predictive Performance of GDF15 and a Standardized Biomarker Index for Abnormal ECG (biomarker analyzed): ROC Analysis and Probability Modelling. Fig. 6a. ROC curve illustrates the diagnostic performance of GDF-15 in predicting biomarker analyzed abnormal ECG findings from both north and south Indian cohort (50 participants) (biological replicates: n = 50; no technical replicates). The curve plots sensitivity against 1 - specificity across different thresholds. The area under the curve (AUC) is 0.8853. Fig. 6b. ROC curve depicting the diagnostic performance of the standardized biomarker index in predicting abnormal ECG findings (50 participants) (biological replicates: n = 50; no technical replicates). The curve plots sensitivity against 1 - specificity across different thresholds. The area under the curve (AUC) is 0.7789. Fig. 6c. Scatter plot depicting the predicted probability of abnormal ECG findings to the standardized biomarker index (50 participants) (biological replicates: n = 50; no technical replicates). The red fitted line is the overall trend, while each point represents an individual prediction.

## Discussion

Our work have utilised computationally sophisticated methods in feature extraction (EMD) and classification (SVM) of ECGs. Multiple studies have also used similar computational methods to classify the ECGs (18–20). Several other deep-learning architectures have been utilised in the ECG classification and prediction of CVDs, such as Convolutional Neural Network (CNN) (21–23), Recurrent Neural Networks (RNN) (24, 25), Gradient Boosting Machine (GBM) (26, 27), and Long Short-Term Memory (LSTM) (28,29). The algorithms validated the classification of normal and abnormal ECG signals with an accuracy of 94% for data based on biomarkers and 90% for ECG data without biomarkers. This result greatly surpasses the performance reported in studies only using ECG data, which usually achieved between 80% and 90% accuracy. For instance, an accuracy of 82.3% using Deep Neural Networks (DNN) (30), 83.7% accuracy using Logistic Regression (31) and 86.6% using Random Forests (32). On the other hand, some computational platforms have shown similar or higher accuracy levels than our study findings, such as 93.6% accuracy using Bidirectional Long Short-Term Memory Networks (33), 94.7% using One-dimensional Convolutional Neural Networks (34) and 95.7% using Generative Adversarial Networks (35). These variabilities might be attributed to the parameters taken to validate the predictive algorithm and the varied population demographics. Adding molecular biomarker data, such as hs-Trop T, GDF-15, and H-FABP, has significantly improved diagnostic accuracy. hs-Trop T was reported to be a reliable marker for MI, showing significantly higher levels among patients having abnormal ECG and MI, irrespective of the regional variations. In contrast, H-FABP showed variation between North and South regions; consistently higher levels were seen among South Indian cohorts than the Northern counterparts, suggesting environmental and socio-demographic influences. Similarly, GDF-15 was elevated among abnormal ECG patients in both regions. However, the North Indian cohort population had lower GDF-15 levels than the South Indian cohort population, indicating regional variations in cardiac stress. The observations made in the paper are a pioneering effort to not only integrate the divergent modalities, including epidemiology, demographic and molecular attributes of the subjects, but also the ECG analyses using AI computational platforms using stable algorithms. The variations in the baseline of the biomarkers in our study, such as H-FABP and GDF-15 between the North and South Indian population, indicate that socio-cultural norms, lifestyle choices and mutation mapping do influence the outcome both in terms of molecular and electrical attributes, which is measured by biomarkers and ECG respectively. Studies with larger populations must extend this work to make this computational platform more amenable.

Although the existing literature has explored the function of AI in ECG classification and biomarkers in CVD detection, integration of these two modalities for CVD detection, considering regional variations, is in a nascent stage. Over the last decade, various AI computational modalities have been studied that use ECG data and other vital sign parameters to predict CVDs but didn’t account for biomarkers (36–48). Similarly, multiple studies have established the diagnostic potential of cardiac biomarkers for detecting CVDs but lacked integration with AI-based ECG analyses (13, 49, 50). Our study bridges these gaps by combining computational ECG analyses with biomarker profiling, offering a holistic diagnostic approach that can outperform traditional methods in accuracy and applicability.

The major strength of our study is the integration of the clinical, molecular, and Intelligent Decision Support System (SVM and EMD approach) to assess cardiovascular risk utilizing the ECG data from the North and South Indian cohorts. The Intelligent Decision Support System-based approach has enhanced the predictive accuracy and interpretability of ECG abnormalities, which forms a scalable platform for early detection of CVDs. This study has considered two distinct geographical populations, which has enabled the identification of region-specific patterns of ECG abnormalities and cardiac biomarker expression, specifically GDF-15. This approach has provided a novel perspective on the predictive utility of the emerging biomarkers which can be based on the geographical distribution. Additionally, standardized ECG annotation and biomarker processing protocols have ensured the data reliability across both the sites. However, this study’s limitation is expressed by the non-representative sampling strategy, as the data was collected from the selected cohorts rather than a nationally representative population. The uneven sample of clinically annotated normal and abnormal ECGs and the limited blood samples from the two geographical cohorts, may affect the generalizability of the outcome. Moreover, the cross-sectional study design limits the casual inferences. The study findings are generalisable to symptomatic adult patients attending tertiary cardiac centres in urban Indian settings. However, applicability may be limited for rural, primary care, or asymptomatic populations. The use of real-world ECGs and biomarker data enhances relevance for similar clinical and diagnostic settings. These setbacks form the rationale for the larger and longitudinal studies using the national representative data from varied geographical centres of India, which will not only validate but also expand our preliminary insights, that can serve as a benchmark. Future efforts should aim to extend this model for broader use in healthcare systems.

## Data Availability

All data produced in the present study are available upon reasonable request to the authors

## Acknowledgement

The authors would like to acknowledge all the patients who participated in the study.

## Author Contribution

DN, KNK and LM collected the data, analyzed the data clinico-epidemiologically and drafted the manuscript. MK, AG and PR have designed and performed the artificially integrated experiments, SM, KMR, and KR did the molecular analysis of the data. SAK performed statistical analysis of the dataset. RK, AO and VED supervised the study and data collection. JS, HS, RD and HS supervised the technical components of the study. RJ designed, oversaw the study, and revised the manuscript. All authors reviewed and edited the manuscript.

