## Supplementary table for "Intelligent Decision Support System Facilitating Early Detection of Cardiovascular Disease"

| | Cardiovascular disease | Diabetes Mellitus | High Cholesterol | High Blood Pressure | Liver Disease |
|---|---|---|---|---|---|
| | n (%) | n (%) | n (%) | n (%) | n (%) |
| **F** | 12(22.2) | 7 (13) | 4(7.4) | 8 (14.8) | - |
| **M** | 2 9(3.7) | 2(3.7) | 2(3.7) | 6(11.1) | 1(1.8) |
| **F+M** | - | 2(3.7) | - | 1(1.8) | - |
| **S** | 4 (7.4) | 6(11.1) | 2(3.7) | 3(5.5) | - |
| **F+M+S** | - | 1(1.8) | - | - | - |
| **M+C** | - | 1(1.8) | - | - | - |
| **C** | - | - | - | - | 1(1.8) |
| **C+S** | - | 1(1.8) | - | 1(1.8) | - |
| **PU** | 1 (1.9) | 1(1.8) | 1(1.8) | - | - |
| **F+S** | 4 (7.4) | - | 2(3.7) | - | - |
| **M+S** | 1 (1.9) | - | 1(1.8) | 2(3.7) | - |
| **F+PU** | - | 1(1.8) | - | 1(1.8) | - |
| **M+F+S+MA+ MU+PA+PU** | - | 1(1.8) | - | - | - |
| **M+S+GP+MU** | - | 1(1.8) | - | - | - |
| **F+GP+PU** | 2 (3.7) | - | 2(3.7) | 1(1.8) | - |
| **M+S+MA+MU** | - | - | - | 1(1.8) | - |

M=Mother, F=Father, S=Sibling, C=Child, GP= Grandparent, MA=Maternal Aunt, MU=Maternal Uncle, PA=Paternal Aunt, PU=Paternal Uncle

Supplementary Table 1 describes the medical history of the patient's family members from South Indian cohort (n-54) (biological replicates: n = 54; no technical replicates). 22.2% of the patient' father had cardiovascular diseases, while 14.8% of the fathers had high blood pressure and 13% of the fathers had diabetes. 11.1% of the patient's mother had high blood pressure, 3.7% of the mothers had cardiovascular diseases and 1.8% had liver diseases.

**Supplementary Table 1. Table describing family medical history of the patients from South Indian cohort (biomarker analyzed)**

| | Father | Mother | Father + Mother |
|---|---|---|---|
| | n (%) | n (%) | n (%) |
| **Family Medical history** | | | |
| Cardiovascular Disease | 4 (12.0) | - | 1(3.0) |
| Diabetes | 3 (9.0) | 4 (12.0) | 1(3.0) |
| High Cholesterol | - | - | 1(3.0) |
| High Blood pressure | 3 (9.0) | 5 (15.0) | 1(3.0) |
| Liver disease | 1(3.0) | 1(3.0) | - |

Supplementary Table 2 describes the medical history of the patient's family members (n-34) (biological replicates: n = 34; no technical replicates). The medical history of cardiovascular disease has been documented among 12% of patients' fathers. 15% of mothers reported high blood pressure.

**Supplementary Table 2. Table describing family medical history of the patients (for molecular analysis) from the North Indian cohort**

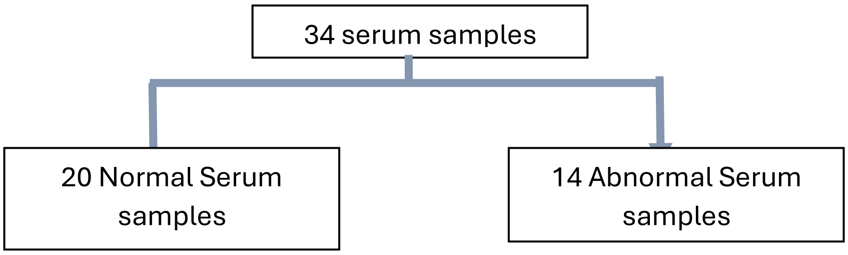


**Supplementary Figure 3. Figure depicting the segregation of serum samples from North Indian cohort for molecular analyses**

The supplementary figure 3 shows the sub-grouping of curated normal and abnormal ECGs serum samples from North Indian cohort for molecular analyses to detect novel biomarkers such as soluble suppression of tumorigenicity-2 (sST2), Heart type Fatty Acid Binding Protein (h-FABP), Growth Differentiation Factor – 15 (GDF-15), High-sensitive Troponin T (hs-Trop T) for predicting impending Cardiovascular (CV) events at both tertiary care centers and their extended community.

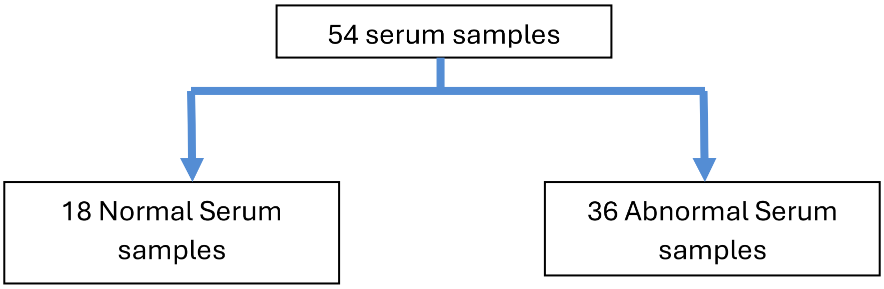

**Supplementary Figure 4. Figure depicting segregation of serum samples from the South Indian cohort for molecular analyses**

The supplementary figure 4 shows the sub-grouping of curated normal and abnormal ECGs serum samples from the South Indian cohort for molecular analyses to detect novel biomarkers such as soluble suppression of tumorigenicity-2 (sST2), Heart type Fatty Acid Binding Protein (h-FABP), Growth Differentiation Factor – 15 (GDF-15), High-sensitive Troponin T (hs-Trop T) for predicting impending Cardiovascular (CV) events at both tertiary care centers and their extended community.

### STROBE Checklist

| | **Item No** | **Recommendation** | **Page No** |
|---|---|---|---|
| **Title and abstract** | 1 | (*a*) Indicate the study's design with a commonly used term in the title or the abstract | 1 |
| | | (*b*) Provide in the abstract an informative and balanced summary of what was done and what was found | 1 |
| **Introduction** | | | |
| Background/rationale | 2 | Explain the scientific background and rationale for the investigation being reported | 2 |
| Objectives | 3 | State specific objectives, including any prespecified hypotheses | 3 |
| **Methods** | | | |
| Study design | 4 | Present key elements of study design early in the paper | 4 |
| Setting | 5 | Describe the setting, locations, and relevant dates, including periods of recruitment, exposure, follow-up, and data collection | 4,5 |
| Participants | 6 | (*a*) Give the eligibility criteria, and the sources and methods of selection of participants | 4 |
| Variables | 7 | Clearly define all outcomes, exposures, predictors, potential confounders, and effect modifiers. Give diagnostic criteria, if applicable | 5-9 |
| Data sources/ measurement | 8* | For each variable of interest, give sources of data and details of methods of assessment | *Source of data-4, details of the method-5-9, Comparability of assessment methods-5-9* |

| | | | |
|---|---|---|---|
| | | (measurement). Describe comparability of assessment methods if there is more than one group | |
| Bias | 9 | Describe any efforts to address potential sources of bias | 4 |
| Study size | 10 | Explain how the study size was arrived at | 5 |
| Quantitative variables | 11 | Explain how quantitative variables were handled in the analyses. If applicable, describe which groupings were chosen and why | 5-9 |
| Statistical methods | 12 | (*a*) Describe all statistical methods, including those used to control for confounding | 8-9 |
| | | (*b*) Describe any methods used to examine subgroups and interactions | 5-9 |
| | | (*c*) Explain how missing data were addressed | 5 |
| | | (*d*) If applicable, describe analytical methods taking account of sampling strategy | 4 |
| | | (*e*) Describe any sensitivity analyses | 7 (Sensitivity analysis in the STROBE context was not applicable, as our study was cross-sectional with a fixed sample. However, robustness of our computational model was evaluated using performance metrics such as accuracy through cross-validation) |
| **Results** | | | |
| Participants | 13* | (a) Report numbers of individuals at each stage of study—eg numbers potentially eligible, examined for eligibility, confirmed eligible, included in the study, completing follow-up, and analysed | 5 & 9 |
| | | (b) Give reasons for non-participation at each stage | 5 (All participants who were approached and met the inclusion criteria consented to participate in the study) |
| | | (c) Consider use of a flow diagram | Supplementary file (figure 3 and 4) |
| Descriptive data | 14* | (a) Give characteristics of study participants (eg demographic, | 10-11 |

| | | | |
|---|---|---|---|
| | | clinical, social) and information on exposures and potential confounders | |
| | | (b) Indicate number of participants with missing data for each variable of interest | - |
| Outcome data | 15* | Report numbers of outcome events or summary measures | Socio-demographic details- 10, Clinical symptoms & medical history- 10 & 11, biomarker's details- 11- 13, ROC curve analyses- 13, Normal and Abnormal ECG status- 5 |
| Main results | 16 | (*a*) Give unadjusted estimates and, if applicable, confounder-adjusted estimates and their precision (eg, 95% confidence interval). Make clear which confounders were adjusted for and why they were included | NA |
| | | (*b*) Report category boundaries when continuous variables were categorized | NA [Biomarkers (e.g., Troponin-T, GDF-15, ST-2) are treated as continuous variables, but we did not categorize them (e.g., no conversion into 'high' vs. 'low' levels)] |
| | | (*c*) If relevant, consider translating estimates of relative risk into absolute risk for a meaningful time period | NA |
| Other analyses | 17 | Report other analyses done—eg analyses of subgroups and interactions, and sensitivity analyses | Subgroup analysis- 11-13, interactions, and sensitivity analyses- NA |
| **Discussion** | | | |
| Key results | 18 | Summarise key results with reference to study objectives | 13-15 |
| Limitations | 19 | Discuss limitations of the study, taking into account sources of potential bias or imprecision. Discuss both direction and magnitude of any potential bias | 16 |
| Interpretation | 20 | Give a cautious overall interpretation of results considering objectives, limitations, multiplicity of analyses, results from similar studies, and other relevant evidence | 13-16 |

| | | | |
|---|---|---|---|
| Generalisability | 21 | Discuss the generalisability (external validity) of the study results | 16 |
| **Other information** | | | |
| Funding | 22 | Give the source of funding and the role of the funders for the present study and, if applicable, for the original study on which the present article is based | 16 |
